# Clinical Features and Predictors for Patients with Severe SARS-CoV-2 Pneumonia: a retrospective multicenter cohort study

**DOI:** 10.1101/2020.06.01.20119032

**Authors:** Chao Cao, Meiping Chen, Yiting Li, Lili Yu, Weina Huang, Guoqing Qian, Chuanbing Zhu, Jinguo Chu, Li He, Jingping Ma, Xiaomin Chen

## Abstract

**Background:** Starting from early December 2019, cases of human infection with a novel coronavirus were identified in Wuhan, Hubei Province, China. It spreads rapidly to other cities and numerous countries. This study was performed to investigate clinical features of patients with severe SARS-CoV-2 pneumonia and identify risk factors for converting to severe cases in those who had mild to moderate diseases.

**Methods:** In this retrospective, multicenter cohort study, patients with mild to moderate SARS-CoV-2 pneumonia were included from Ningbo First Hospital and Jingzhou Central Hospital. Demographic data, symptoms, laboratory values, comorbidities, and clinical outcomes were collected. Data were compared between non-severe and severe patients. Logistic regression analysis was performed to assess risk factors in predicting the patients with SARS-CoV-2 pneumonia who would convert to severe cases.

**Findings:** 120 patients (36 from Ningbo First Hospital and 84 from Jingzhou Central Hospital) were included in this study, among which 62 were excluded and 58 were included in the final analysis. Compared with non-severe cases, severe patients with SARS-CoV-2 pneumonia had a longer: time to clinical recovery (12.9±4.4 *vs* 8.3±4.7; p=0.0011), duration of viral shedding (15.7±6.7 *vs* 11.8±5.0; p=0.0183), and hospital stay (20.7±1.2 *vs* 14.4±4.3; p=0.0211). Multivariate logistic regression indicated that lymphocyte count was significantly associated with the rate of converting to severe cases (odds ratio 1.28, 95%CI 1.06–1.54, per 0.1×10^9^/L reduced; p=0.007), while using of low-to-moderate doses of systematic corticosteroids was associated with reduced likelihood of converting to a severe case (odds ratio 0.14, 95%CI 0.02–0.80; p=0.0275).

**Interpretation:** The low peripheral blood lymphocyte count was an independent risk factor for SARS-CoV-2 pneumonia patients converting to severe cases. This finding may help clinicians more accurately predict prognosis, and triage priorities to improve clinical outcomes.

**Research in context:** *Evidence before this study:* Severe Acute Respiratory Syndrome Coronavirus 2 (SARS-CoV2) is a novel coronavirus that have emerged in early December 2019, and has caused a novel coronavirus disease (COVID-19). It has been deemed as a public health emergency of global concern by The World Health Organization (WHO). We searched PubMed for articles published up to March 11, 2020, using the search terms (“novel coronavirus” OR “SARS-CoV-2” OR “COVID-19”) with no language or time restrictions. Previous work has described clinical characteristics of critically ill and non-critically ill patients with COVID-19. However, no published works have focused on clinical features of patients with severe SARS-CoV-2 pneumonia and identify risk factors for converting to severe cases in those who had mild to moderate diseases.

*Added value of this study:* In this retrospective and multicenter cohort study, we reported demographics characteristics, baseline symptoms, laboratory findings, corticosteroid usage and hospital course of patients with non-severe COVID-19 and severe COVID-19. Comparing with non-severe patients, severe patients with COVID-19 was found to have a longer: time to clinical recovery (12.9±4.4 *vs* 8.3±4.7; p=0.0011), duration of viral shedding (15.7±6.7 *vs* 11.8±5.0; p=0.0183), and hospital stay (20.7±1.2 *vs* 14.4±4.3; p=0.0211). By multivariate logistic regression, we found increasing odds of converting to severe cases associated with lower lymphocyte count (odds ratio 1.28, 95%CI 1.06–1.54, per 0.1×10^9^/L reduced; p=0.007). Using of low-to-moderate doses of systematic corticosteroids was associated with reduced likelihood of converting to a severe case (odds ratio 0.14, 95%CI 0.02–0.80; p=0.0275).

*Implications of all the available evidence:* Low lymphocyte count in peripheral blood was an independent risk factor for patients who converted to severe cases. In addition, using of systematic corticosteroids in mild to moderate patients with SARS-CoV-2 pneumonia was associated with a reduced risk of converting to severe cases. These findings may help clinicians predict prognosis more accurately, and triage priorities to improve clinical outcomes. Further prospective studies are warranted to confirm these findings.

## Introduction

Starting from early December 2019, cases of human infection with a novel coronavirus were identified in Wuhan, Hubei Province, China. It spreads rapidly to other cities and numerous countries by human-to-human transmission^1–3^. This disease has been deemed as a public health emergency of global concern by The World Health Organization (WHO) and named as coronavirus infected disease 2019 (COVID-19)^4^. Confirmed by high-throughput sequencing, COVID-19 is caused by Severe Acute Respiratory Syndrome Coronavirus 2 (SARS-CoV-2), a novel beta-coronavirus, showing around 79% identity with Severe Acute Respiratory Syndrome Coronavirus (SARS-CoV) and 50% identity with Middle East Respiratory Syndrome coronavirus (MERS-CoV)^5^. It is usually accompanied by pneumonia, fever, non-productive cough and asthenia^6^. Nausea, vomiting, and dyspnea are also found in patients with COVID-19, though these are less common^7^. Patients with severe disease may develop Acute Respiratory Distress Syndrome (ARDS), acute cardiac injury, shock and can require invasive ventilation^8^.

Up to now, large numbers of patients with SARS-CoV-2 pneumonia had developed into severe cases or even resulted in death^4^. Guan et al reported findings of 1099 cases with SARS-CoV-2 infection from 552 hospitals in 30 provinces of China and results suggested that 15.7% of patients had developed severe illness and 1.4% of patients died^6^. Among those severe cases, 43 patients (24.9%) were either admitted to ICU, required mechanical ventilation, or died^6^. The median time from onset of symptoms to shortness of breath, an indicator of severe SARS-CoV-2 pneumonia, was 8 days^7^. It was reported that older patients with comorbidities were more likely to develop severe disease^6,8^. However, not all older patients with comorbidities convert to severe cases, indicating that there may be other risk factors. Given the rapid spread of SARS-CoV-2 pneumonia and the large threat to public health, this study aims to investigate clinical features and laboratory findings between non-severe and severe patients, thereby exploring predictive factors for severe cases.

## Methods

### Study design and patient populations

This retrospective analysis was conducted using data collected between Jan 21, 2020 and Feb 25, 2020 from Ningbo First Hospital and Jingzhou Central Hospital. Patients with laboratory confirmed diagnoses of SARS-CoV-2 and had been admitted to one of the above medical centers were included. All patients were identified by positive results of reverse transcriptase polymerase chain reaction (RT-PCR) on SARS-CoV-2 in lower respiratory specimens. Patients enrolled presented with mild to moderate SARS-CoV-2 pneumonia at baseline. To avoid potential effects of antiviral drugs, the largest cohort of patients were selected for analysis; all were initiated on combination therapy of arbidol and Kaletra. Ethical approvals for this study were obtained from the Ethics Commission of Ningbo First Hospital (2020-R017) and the Ethics Commission of Jingzhou Central Hospital (2020–2–19). Written informed consent was waived due to the rapid emergence of this disease.

### Data collection

Patients’ electronic medical records were reviewed to extract the following information retrospectively. The following data were collected: Demographic characteristics: age, gender and history of chronic disease; Symptoms: temperature, cough, dyspnea, score throat, muscle soreness, fatigue, anorexic, nausea and diarrhea; Laboratory assessments on admission: white blood cell count, peripheral blood lymphocyte (PBL), D-dimer, Alanine aminotransferase (ALT), Aspartate Aminotransferase (AST) and C-reactive protein (CRP); Dates of important events: onset of symptoms, onset of antiviral therapy, admission to hospital time, discharge or death and the date of negative result of viral nucleic acid test; Details of therapeutic regimen: antiviral medicines and use of corticosteroids.

### Definitions

The severity scores were calculated within 48h of hospital admission. Patients were divided into two cohorts according to the version of Diagnosis and Treatment of Pneumonia Caused by COVID-19 (version 6) issued by National Health Commission of the People’s Republic of China^9^. Cohort 1, patients who did not convert to severe cases; Cohort 2, patients who converted to severe cases. Patients who met at least one of the following criteria were defined as the severe cases^9^: (1). Respiratory rate ≥ 33/minute; (2). Oxygen saturation ≤ 94% in resting state; (3). PO2/FiO2 ≤ 300mmHg; (4). Developed respiratory failure and require mechanical ventilation; (5). Developed shock; (6). Multiple organ dysfunction and requiring ICU care. The duration (in days) from the start of antiviral therapy to clinical recovery was selected as the primary outcome measure. Clinical recovery is defined as the remission of clinical symptoms for at least 72 hours, including normalization of respiratory rate (≤24/minute on room ≤ air), oxygen saturation (>94% on room air), fever (≥37°C of axilla, ≥37.2 °C of oral) and cough (no cough or mild cough).

### Statistical Analysis

Continuous variables were expressed as mean (SD) or median (IQR) and categorical variables as quantity (proportion). The t-test or Mann-Whitney *U* test was used for comparison of continuous variables. The chi-squared (χ2) test was used to compare categorical variables. Logistic regression was performed to assess the usefulness of risk factors in predicting the patients with mild to moderate SARS-CoV-2 pneumonia who would convert to severe cases. The following factors were analyzed to determine their effects: age, sex, fever, time from onset of symptoms to antiviral treatment, comorbidities, CRP, PBL, and use of corticosteroids. Odds Ratios (ORs) and 95% confidence intervals (CIs) were estimated. A 2-sided α of <0.05 was considered statistically significant. Statistical analyses were conducted using SPSS version 180 (SPSS, Chicago, IL, USA) and SAS version 9.4 (SAS Institute, Cary, North Carolina).

## Results

120 patients with SARS-CoV-2 pneumonia had been admitted to Ningbo First Hospital and Jingzhou Central Hospital, among which 58 patients with mild to moderate disease who received the same antiviral drugs were included (figure 1). The mean age (SD) of patients was 47.8 years (13.8), where patients aged 40–79 years accounted for 63.8% (table 1). 30 (51.7%) patients were male. 15 (25.9%) cases had underlying chronic diseases, and 9 (15.5%) had hypertension. Cough (58.6%), fever (41.4%) and fatigue (31.0%) were the most common symptoms at onset, followed by muscle soreness (17.2%) and dyspnea (15.5%). A higher prevalence of comorbidities (34.8% *vs* 20.0%) was observed in patients who converted to severe cases, and more male patients also converted to severe cases (73.9% *vs* 37.1%). 25 (43.1%) patients received methylprednisolone 1–2 mg/kg per day.

**Figure 1:**
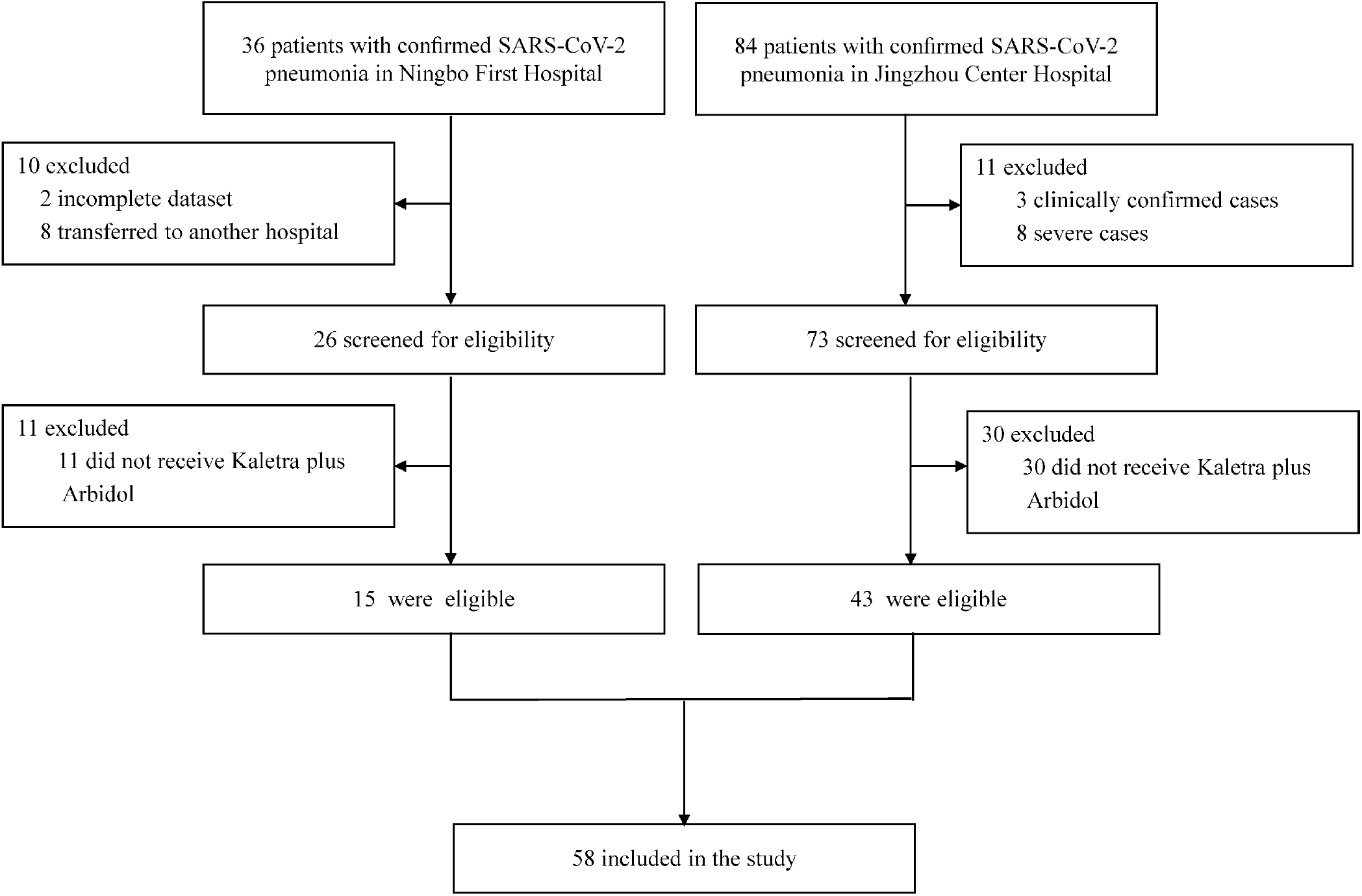
Flowchart of enrolled patients.

**Table 1:** Demographics and baseline characteristics of patients with SARS-CoV-2 pneumonia.

|  | **All patients**<br>**(n=58)** | **Non-severe**<br>**(n=35)** | **Severe**<br>**(n=23)** |
| --- | --- | --- | --- |
| **Age, years** | 47.8 (13.8) | 46.0 (12.7) | 50.5 (15.2) |
| **Age range, years** |  |  |  |
| 20-29 | 7 (12.1%) | 5 (14.3%) | 2 (8.7%) |
| 30-39 | 13 (22.4%) | 8 (22.9%) | 5 (21.7%) |
| 40-49 | 9 (15.5%) | 6 (17.1%) | 3 (13.0%) |
| 50-59 | 19 (32.8%) | 11 (31.4%) | 8 (34.8%) |
| 60-69 | 7 (12.1%) | 5 (14.3%) | 2 (8.7%) |
| 70-79 | 2 (3.5%) | 0 | 2 (8.7%) |
| ≥80 | 1 (1.7%) | 0 | 1 (4.4%) |
| **Gender** |  |  |  |
| Female | 28 (48.3%) | 22 (62.9%) | 6 (26.1%) |
| Male | 30 (51.7%) | 13 (37.1%) | 17 (73.9%) |
| **Comorbidities** |  |  |  |
| None | 43 (74.1%) | 28 (80.0%) | 15 (65.2%) |
| Hypertension | 9 (15.5%) | 4 (11.4%) | 5 (21.7%) |
| Diabetes | 1 (1.7%) | 0 | 1 (4.4%) |
| Chronic kidney disease | 1 (1.7%) | 0 | 1 (4.4%) |
| Malignancy | 1 (1.7%) | 0 | 1 (4.4%) |
| Others | 5 (8.6%) | 3 (8.6%) | 2 (8.7%) |
| **Symptoms** |  |  |  |
| Fever | 24 (41.4%) | 12 (34.3%) | 12 (52.2%) |
| Cough | 34 (58.6%) | 22 (62.9%) | 12 (52.2%) |
| Sore throat | 6 (10.3%) | 2 (5.7%) | 4 (17.4%) |
| Nausea | 6 (10.3%) | 5 (14.3%) | 1 (4.4%) |
| Anorexic | 5 (8.6%) | 4 (11.4%) | 1 (4.4%) |
| Diarrhea | 4 (6.9%) | 3 (8.6%) | 1 (4.4%) |
| Muscle soreness | 10 (17.2%) | 6 (17.1%) | 4 (17.4%) |
| Dyspnea | 9 (15.5%) | 4 (18.2%) | 5 (23.8%) |
| Fatigue | 18 (31.0%) | 10 (28.6%) | 8 (34.8%) |
| **Severity of cough** |  |  |  |
| Absent | 24 (41.4%) | 13 (37.1%) | 11 (47.8%) |
| Mild | 10 (17.2%) | 9 (25.7%) | 1 (4.4%) |
| Moderate | 18 (31.0%) | 10 (28.6%) | 8 (34.8%) |
| Severe | 6 (10.3%) | 3 (8.6%) | 3 (13.0%) |

Data are n (%), mean (SD) or median (IQR), unless otherwise stated.

Time from onset of illness to antiviral treatment for Cohort 1 was 3 (IQR 1–7) days and 4 (IQR 2–6) days in Cohort 2 (table 2). Patients who converted to severe cases were associated with having a lower lymphocyte count (Cohort 1: 1.3±0.4, *vs* Cohort 2: 0.9±0.4, *P* = 0.0064) and a higher CRP (Cohort 1: 4.5 [0.6–10.9] *vs* Cohort 2: 16.8[3.9–55.8], *P* = 0.0041) on admission. Compared with non-severe cases, severe patients had a longer time to clinical recovery (Cohort 1: 8.3±4.7 *vs* Cohort 2: 12.9±4.4, *P* = 0.0011), duration to negative viral nucleic acid test (Cohort 1: 11.8±5.0 *vs* Cohort 2: 15.7±6.7, *P* = 0.0183), and hospital stay (Cohort 1: 14.4±4.3 *vs* Cohort 2: 20.7±1.2, *P* = 0.0211).

**Table 2:** Laboratory findings, corticosteroid usage, and hospital course in patients with SARS-CoV-2 pneumonia.

|  | **Non-severe**<br>**(n=35)** | **Severe**<br>**(n=23)** | ***p-value*** |
| --- | --- | --- | --- |
| **Laboratory findings** |  |  |  |
| WBCs count, $\times 10^9$ /L | 5.1 (1.9) | 5.9 (4.1) | 0.3 |
| Lymphocyte count, $\times 10^9$ /L | 1.3 (0.4) | 0.9 (0.4) | 0.0 |
| C-reactive protein, mg/L | 4.5 (0.6-10.9) | 16.8 (3.9-55.8) | 0.0 |
| D-dimer, mg/L | 90.0 (45.5-180.0) | 154.0 (113.3-470.5) | 0.1 |
| ALT level, IU/L | 20.9 (8.9) | 28.3 (22.4) | 0.1 |
| AST level, IU/L | 24.0 (6.3) | 31.0 (13.3) | 0.0 |
| **Corticosteroid usage** |  |  | 0.0 |
| Yes | 11 (31.4%) | 14 (60.9%) |  |
| No | 24 (68.6%) | 9 (39.1%) |  |
| **Hospital course** |  |  |  |
| Time from onset of illness to antiviral treatment, days | 3.0 (1.0-7.0) | 4.0 (2.0-6.0) | 0.6 |
| Clinical recovery time, days | 8.3 (4.7) | 12.9 (4.4) | 0.0 |
| Time of virus nucleic acid turn to negative, days | 11.8 (5.0) | 15.7 (6.7) | 0.0 |
| Hospitalization duration, days | 14.4 (4.3) | 20.7 (1.2) | 0.0 |

Abbreviations: WBCs, white blood cell counts; ALT, Alanine aminotransferase; AST, Aspartate Aminotransferase.

Multivariate logistic regression indicated that lymphopenia (peripheral blood lymphocyte count < 1.1×10^9^/L) is an independent risk factors for patients converting to severe cases, with OR value of 8.0 (95%CI, 1.5–41.8) converted to severe cases. The OR for converting to severe cases was 1.28 (95%CI, 1.06–1.54) in patients with a reduced of 0.1×10^9^/L peripheral blood lymphocyte (table 3). In addition, the OR for use of corticosteroids in mild to moderate patients with SARS-CoV-2 pneumonia was 0.14 (95%CI, 0.02–0.80) in those who converted to severe cases.

**Table 3:** Logistic regression assessed the risk factors in predicting patients with mild to moderate SARS-CoV-2 pneumonia conversion rate to severe cases.

| Variable | Regression<br>coefficient | Standard<br>error | *P* value | Odds<br>ratio | Lower<br>OR | Upper<br>OR |
| --- | --- | --- | --- | --- | --- | --- |
| Age, years | -0.04 | 0.04 | 0.2376 | 0.96 | 0.90 | 1.03 |
| Gender (female vs. male) | -1.38 | 0.76 | 0.0702 | 0.25 | 0.06 | 1.12 |
| Fever (yes vs. no) | 1.13 | 0.60 | 0.0597 | 3.09 | 0.96 | 9.99 |
| Comorbidities (yes vs. no) | 0.87 | 0.86 | 0.3121 | 2.38 | 0.44 | 12.76 |
| C-reactive protein (<10 mg/L vs. ≥10 mg/L) | 0.38 | 0.75 | 0.6145 | 1.46 | 0.33 | 6.40 |
| Lymphocyte count (per reduced $0.1 \times 10^9$/L) | -0.25 | 0.09 | 0.0070 | 0.78 | 0.65 | 0.94 |
| Time from onset of illness to antiviral treatment,<br>days | 0.03 | 0.06 | 0.6670 | 1.03 | 0.91 | 1.17 |
| Corticosteroids (yes vs. no) | -1.99 | 0.90 | 0.0275 | 0.14 | 0.02 | 0.80 |

## Discussion

The most common symptoms for SARS-CoV-2 pneumonia at onset of illness were fever and cough, but some patients had presence of dyspnea at a median of 8 days after onset of illness^9^. Dyspnea, usually observed in severe patients with SARS-CoV-2 pneumonia^10^, may indicate the progression of disease with a low oxygenation index and a state of severe illness. However, up to now, the clinical course of SARS-CoV-2 pneumonia and risk factors for converting to severe cases remain unknown.

This study was performed to investigate clinical characteristics of patients with severe SARS-CoV-2 pneumonia, and identify risk factors for those with mild to moderate disease who converted to severe cases. 120 patients (36 from Ningbo First Hospital and 84 from Jingzhou Central Hospital) were included in this study, among which 62 were excluded and 58 were included in the final analysis. The results from this study showed that low peripheral blood lymphocyte count (< 1.1×10^9^/L) was an independent risk factor for patients converting to severe cases (OR: 8.0, 95%CI: 1.5–41.8). With each reduction of 0.1×10^9^/L in peripheral blood lymphocyte count, the OR for converting to severe cases increased 28% (95%CI, 6%–54%). In addition, using of corticosteroids in mild to moderate patients with SARS-CoV-2 pneumonia was also associated with a reduced risk of converting to severe cases (OR: 0.14, 95%CI: 0.02–0.80).

Lymphopenia was also observed in most patients with SARS-CoV infection during their course of illness^11,12^. He et al believed that lymphopenia was a significant factor of SARS-CoV infection and lymphocyte counts may be useful in predicting the disease severity and clinical outcomes^12^. A recent study reported an absolute lymphopenia (lymphocyte count < 1.0×10^9^/L) in 63% of patients with SARS-CoV-2 pneumonia on admission. Low levels of total lymphocyte counts were more marked in those patients in which infection resulted in death^10^. A possible cause for the lymphopenia may be that lymphocytes are directly infected and destroyed by SARS-CoV-2. However, this requires further study to confirm, as the cellular receptor for SARS-CoV-2 is angiotensin-converting enzyme 2^13^, which is not expressed on B or T lymphocytes^14^. Therefore, depletion of lymphocytes may be secondary to the direct effect of the virus on the lymphocytes or the effect of various cytokine mediated altered lymphocyte trafficking.

Since no antiviral treatment for SARS-CoV-2 infection has been proven to be effective, management of this disease remains clinically based and supportive^15^. Considering that acute viral pneumonia is an important cause of acute lung injury (ALI), the value of systematic corticosteroids in patients with SARS-CoV-2 infection is a focus of interest. This study showed patients who received low-to-moderate dose of systematic corticosteroids were less likely to convert to severe cases. In a report by Chen et al^16^, the use of systematic corticosteroids in SARS resulted in reduced mortality and a shorter hospitalization stay, and was not associated with significant secondary lower respiratory tract infection or other complications. However, it is well known that corticosteroid therapy is a double-edged sword^17,18^. The immune response may be weak and consequently have more difficulty in eradicating the virus because of treatment with systematic corticosteroids^19^. Further studies are urgently required to assess whether systematic corticosteroid treatment is beneficial in patients with SARS-CoV-2 pneumonia.

Some limitations of this study should be acknowledged. Firstly, this was a retrospective study with a limited sample size, if the conclusion generalized is to be widely used, it would still need a prospective large-scale clinical validation. Secondly, different varieties of traditional Chinese medicine were administered to most patients, and the effectiveness and potential adverse effects of those drugs given remain largely unknown. Thirdly, examinations of lymphocyte subsets were undertaken in this study, and in addition, total lymphocyte counts were not observed dynamically.

Despite the above limitations, we believe that our study has shown important and novel findings about the predictors of severe cases in mild to moderate patients with SARS-CoV-2 pneumonia. To our knowledge, this is the first study that has demonstrated that lymphocyte count is a useful predictor for severity of SARS-CoV-2 pneumonia. This may help clinicians more accurately predict prognosis, and triage priorities to improve clinical outcomes.

The findings from this study showed that low lymphocyte count (< 1.1×10^9^/L) in peripheral blood was an independent risk factor for patients who converted to severe cases. In addition, using of systematic corticosteroids in mild to moderate patients with SARS-CoV-2 pneumonia was associated with a reduced risk of converting to severe cases. Further prospective studies are warranted to confirm these findings.

## Data Availability

Participant data without names and identifiers will be made available after approval from the corresponding author.

## Contributors

CC, LH, JM, LY, and CZ collected the epidemiological and clinical data. CC and MC processed statistical data. YL, WH, GQ, JC, and XC summarized all data. CC and JM drafted the manuscript, CC, LH and XC reviewed the final manuscript.

## Acknowledgments

We thank Phoebe Jaye Miles for writing assistance and revised of the manuscript.

